# Creating a Survey Instrument for Self-Assessed Menstrual Cycle Characteristics and Androgen Excess

**DOI:** 10.1101/2020.03.03.20030676

**Authors:** Shruthi Mahalingaiah, Carol Cosenza, J. Jojo Cheng, Erika Rodriguez, Ann Aschengrau

**Author notes:** **Corresponding author:** Shruthi Mahalingaiah. **Reprint Requests:** Shruthi Mahalingaiah, Department of Environmental Health, Harvard T.H. Chan School of Public Health, Building 1 665 Huntington Avenue, Boston, MA 02115, USA. **Acknowledgements** SM would like to acknowledge that much of this work was performed at Boston University Medical Campus (BUMC). SM would also like to acknowledge the Departments of OB/GYN at BUMC for their care of diverse populations. SM also acknowledges Barbara Corkey, PhD, who provided mentorship that was critical to build the research team and Renee Cannon for illustrating the images in this survey and for granting one-time US right re-use for journal publication. **Contributorship Statement** SM conceived the idea for the project, searched for the literature to create questions, and managed the collaborations. She wrote the first draft of the paper and revised several drafts before approving the final version. She agrees to be accountable for all aspects of the work. CC conducted the cognitive testing and reviewed several manuscript drafts. JJC searched the literature for questions, revised survey versions, and revised several manuscript drafts. ER conducted literature searches, assisted on the first draft, revised manuscript drafts, and prepared the manuscript for submission. AA informed the process of cohort development and revised drafts.

## Abstract

Polycystic ovary syndrome (PCOS) is the most common reproductive disorder in women, but tools for self-reported ascertainment are currently lacking. We created and evaluated an online survey instrument for identification of PCOS by incorporating clinically derived reproductive health questions and a novel pictorial tool for androgen excess. The survey was cognitively tested for question comprehension and usability. Comprehension problems included simplifying the PCOS definition and questions on bleeding duration. Recall and answer formation problems included beverage consumption and questions using symbols. Evaluation of this survey instrument via cognitive testing may help to accurately ascertain the prevalence of PCOS in general population settings.

## Introduction

Polycystic ovary syndrome (PCOS) is the most common endocrine disorder in reproductive aged women and is characterized by rare ovulation and associated menstrual irregularity, androgen excess, and in some cases, presence of polycystic ovarian morphology on ultrasonographic or other radiologic assessments (Sheehan, 2004). Women with PCOS are at increased risk for endometrial hyperplasia (Charalampakis et al., 2016), infertility (Aziz et al., 2012), and metabolic disorders including diabetes, dyslipidemia (Legro et al., 2013), obesity (Flegal et al., 2010), and hypertension (Wild et al., 2000). Women with PCOS are also at increased risk for mood disorders (Legro et al., 2013), such as depression and anxiety, and non-alcoholic fatty liver (Cerda et al., 2007).

Ascertainment of PCOS, menstrual cycle characteristics, and androgen excess have been limited in large population-based epidemiologic studies. Often, surveys ask a single question about having been diagnosed with PCOS or use self-reported menstrual irregularity as a proxy for PCOS status (Chavarro et al., 2016; Mahalingaiah et al., 2017; Mahalingaiah et al., 2016). Because menstrual disorders, such as PCOS, often require a clinical diagnosis for respondents to be able to report their disease status, mild or clinically unrecognized disease are often not captured in population-based studies. Existing tools are also limited in how to quantify the clinical characteristics of androgen excess by self-report, including hirsutism, acne, and alopecia. Our goal was to design a survey instrument for use in an online survey that would: 1) ascertain self-reported menstrual history and irregular menstrual cycle phenotypes and 2) capture self-reported androgen excess including hirsutism, acne, and alopecia that could be used in a diverse population of women. This report describes the process of survey design and evaluation via cognitive testing that was undertaken to create the survey for The Ovulation and Menstruation Health (OM) Pilot Study.

## Methods

The survey instrument was designed in two phases. During the first phase, the questionnaire including the survey questions and the pictorial self-assessment tools were created. In the second phase, an expert review of the draft questionnaire and pictorial tools was conducted. Cognitive testing was conducted using a print version for comprehension. Usability testing was conducted using the online survey instrument.

### Phase 1

#### Questionnaire Creation

The design process began with a review of existing questionnaires from women’s health studies such as the Nurses’ Health Study 2 (NHS2), the Framingham Heart Study (FHS), and the Cape Cod Health Study (CCHS). These are large, prospective or reproductive cohorts with surveys that span a long temporal window. Table 1 identifies the cohorts used in the initial questionnaire review and whether their questionnaire is publicly available. Although none of these questionnaires were designed to specifically capture menstrual characteristics or other features of androgen excess required to diagnose PCOS, they served as a basis for writing our questions. Additionally, diagnostic criteria of PCOS by the Rotterdam Criteria (Group, 2004) and classification of abnormal uterine bleeding during a woman’s reproductive years (Fraser et al., 2011) by the International Federation of Gynecology and Obstetrics (FIGO) were used as references in drafting questions. A total of 219 questions were developed, which does not reflect the total number of questions an individual may answer due to skip patterns.

**Table 1.** Select cohorts for initial review of women’s health questions with questionnaire availability status.

| Cohort | Publicly Available Questionnaire (Yes/No) |
| --- | --- |
| Nurses’ Health Study 2<sup>a</sup> | Yes |
| Framingham Heart Study<sup>b</sup> | Yes |
| Cape Cod Health Study<sup>c</sup> | No |
| Genes for Good<sup>d</sup> | Yes |
| Women’s Health Initiative<sup>e</sup> | Yes |
| Black Women’s Health Study<sup>f</sup> | Yes |
| Growing Up Today Study<sup>g</sup> | Yes |

<sup>a</sup>Questionnaires: Nurses' Health Study. Brigham and Women's Hospital, Harvard Medical School, Harvard T.H. Chan School of Public Health. https://www.nurseshealthstudy.org/participants/questionnaires. Published 2016. Accessed.
<sup>b</sup>FHS Exam Forms. The Framingham Heart Study: Boston University and the National Heart, Lung, and Blood Institute. https://www.framinghamheartstudy.org/fhs-for-researchers/list-of-exam-forms/. Published 2020. Accessed.
<sup>c</sup>Mahalingaiah S, Winter MR, Aschengrau A. Association of prenatal and early life exposure to tetrachloroethylene (PCE) with polycystic ovary syndrome and other reproductive disorders in the cape cod health study: A retrospective cohort study. *Reprod Toxicol*. 2016;65:87-94.
<sup>d</sup>Genes for Good: For Researchers. University of Michigan: School of Public Health. https://genesforgood.sph.umich.edu/about_study/for_researchers. Published 2019. Accessed.
<sup>e</sup>Women's Health Initiative Forms. Women's Health Initiative. https://www.whi.org/researchers/studydoc/SitePages/Forms.aspx. Accessed.
<sup>f</sup>Black Women's Health Study Questionnaires. Boston University Slone Epidemiology Center. https://www.bu.edu/bwhs/for-researchers/sample-bwhs-questionnaires/. Accessed.

#### Pictorial Self-assessment tool creation

The purpose of creating pictorial tools was to make it easier for respondents to answer questions about their body shape, acne, hirsutism, and alopecia by providing visual examples of the different response categories (Mahalingaiah, 2020). The Ferriman-Gallwey (F-G) score was created in 1961 and has been used to measure hirsutism as a marker of androgen excess in women (Ferriman & Gallwey, 1961). Originally created to assess eleven body areas, the scoring system was later modified to evaluate nine key areas associated with hormonal hair growth. The modified Ferriman-Gallwey (mFG) pictorial tool scores hair growth from no terminal growth to severe levels of growth (0 to 4, for a maximum score of 36). Because different races/ethnicities may have different manifestations of androgen excess, including acne and alopecia, we expanded androgen excess ascertainment to encompass this variation (Cheewadhanaraks et al., 2004; DeUgarte et al., 2006; Yildiz et al., 2010). A medical illustrator was commissioned to sketch the mFG grading scale based on her previous work illustrating the F-G scale (Bode et al., 2012). Images for varying stages of androgenic alopecia based on the Sinclair system were also created (Gupta & Mysore, 2016). Images for acne severity were illustrated corresponding to acne categories within the question stem. Lastly, anthropometric images were created for use in body shape self-report as a supplement to survey questions on body shape.

#### Phase 2: Expert Review and Cognitive Testing

After drafting the survey instrument, the Center for Survey Research (CSR) at the University of Massachusetts Boston was contracted to conduct an expert review on the design, formatting, and usability of the survey instrument (Fowler, 2014; Fowler, 1995). After this review, the survey instrument was built into an online platform using REDCap. The CSR then conducted cognitive interviews for understanding participant comprehension and online survey usability.

### Cognitive Testing

#### Participant Selection for Cognitive Testing

Because the survey instrument was intended to be accessible to an educationally and racial/ethnically diverse population, we conducted in-person interviews in English with six women, aged 22-46 who represented a mix of racial and ethnic identities as well as varying levels of educational attainment (Table 2). Half of the respondents had received a diagnosis of PCOS from a health care provider.

**Table 2.** Demographics of six cognitive testing respondents

|  |  |
| --- | --- |
| **Age (years)** |  |
| Range | 22-46 |
| Mean (SD) | 32.6 (9.2) |
| Median (25<sup>th</sup> Percentile, 75<sup>th</sup> Percentile) | 30 (27, 39) |
| **Race/Ethnicity** |  |
| White | 1 |
| Hispanic | 2 |
| Black or African American | 1 |
| More than 1 Race/Ethnicity | 2 |
| **Education** |  |
| Some College | 4 |
| 4-year College Degree | 2 |
| **PCOS Status** |  |
| Has PCOS | 3 |
| Does not have PCOS | 3 |
| **Number of Prior Pregnancies** |  |
| Range | 0-9 |
| Mean (SD) | 2.5 (3.5) |
| Median (25<sup>th</sup> Percentile, 75<sup>th</sup> Percentile) | 2 (0, 3.2) |
| **Number of Prior Live Births** |  |
| Range | 0-4 |
| Mean (SD) | 1.2 (1.8) |
| Median (25<sup>th</sup> Percentile, 75<sup>th</sup> Percentile) | 0 (0, 2.25) |

#### Cognitive Testing of Survey Instrument

The cognitive testing protocol was designed by CSR and was considered exempt by the IRB at the University of Massachusetts Boston. Interviews were conducted at CSR’s office by two experienced interviewers. Interviewers followed a semi-structured protocol that began with respondents filling out sections of the survey alone. Then, the interviewer reviewed the questions with the respondent by asking a series of follow-up probing questions to gain an understanding of how the survey questions were understood and how respondents formed their answers. The cognitive interviews lasted between 50 to 90 minutes and respondents were paid $80 for their participation. Based on the participants’ feedback, changes were made to the survey instrument to improve its clarity.

Because the full survey was long, we focused the cognitive testing on a limited number of key questions. Particularly important were questions regarding menstrual cycle length, age at menarche, menstrual flow, and body hair growth, as these are the main self-reported characteristics used to ascertain PCOS. Additional questions that were evaluated included those critical in our future analyses such as demographics, anthropometrics, PCOS diagnosis, general health, lifestyle and diet, and pregnancy history. Of the 219 questions created, 120 questions were cognitively tested. We present 14 examples here.

## Results

Cognitive testing allowed us to evaluate how respondents handled each step of the question-answering process. We found issues with <u>comprehension and readability</u> (including formatting), <u>recall and answer formation problems</u>, and <u>usability concerns</u>. This section describes select examples of some of these problems and how we attempted to solve them. The full cognitive testing report by CSR entitled *Ovulation and Menstruation Health Study, Report on Cognitive Testing, July 2017*, stored on Harvard Dataverse. (*Mahalingaiah, 2020)*.

**<u>Comprehension and Readability Problems</u>** include questions that were missing a needed definition, questions that have a definition that is too complex or unclear, questions that are formatted in a way that respondents understand the question in an inconsistent manner, and questions that have embedded assumptions.

### Example 1: Defining Polycystic Ovary Syndrome

Survey Question: Polycystic Ovary Syndrome is a health condition where a woman has fewer than 8 periods each year. It is also associated with increased hair growth on the body. Some women may also have many cysts on the ovaries. Has a doctor ever diagnosed you with Polycystic Ovary Syndrome or PCOS?

Findings: Although this question included a definition, respondents were still confused, including those who had been diagnosed with PCOS. Because the definition was very specific, respondents were unsure how to answer if they did not meet all of the requirements of the definition. The definition was modified in future versions to read: *Polycystic Ovary Syndrome is a health condition involving irregular periods, excess testosterone, increased acne, body and facial hair, and many small cysts in the ovaries. Some women also experience hair loss on the scalp*.

### Example 2: Historic Menstrual Regularity

Survey Question: In the past couple of years, has there ever been a time when your menstrual period was NOT regular or predictable for more than a 3-month window of time?

Findings: Having more than one time-frame in the question (“past couple of years” and “3-month window”) was confusing to respondents who seemed either to answer about one time-frame or the other. The introductory phrase “in the past couple of years” was removed in the final version of the questionnaire.

### Example 3: Menstrual Cycle-Duration of Bleeding

Survey Question: In the last year, when you had your period, about how many days did it usually last? Please only count the days when you were bleeding, not when you were spotting.

Finding: For most people, a question mark signifies the end of a question – at which point it should be answered. However, in this example, important information was provided in the explanatory sentence <u>after</u> the question. Several respondents were unsure whether or not to include days when they were spotting. When asked about it, respondents said they did not notice the second sentence and simply stopped reading at the question. The question was changed to: “Now we want to know about how long your period usually lasts. Counting only the days when you were bleeding and not when you were spotting, in the last year about how many days did your period usually last?”

### Example 4: Hair Removal Frequency

Survey Question: In the last year, how often did you use any hair removal method on any area of your body other than underarms or legs? Response options: Daily, three times a week, twice a week, weekly, twice a month, monthly

Finding: This question assumes a consistency of behavior that may not exist. Respondents were unsure how to answer when hair removal was not on a regular schedule, when they removed hair from different body parts on a different schedule, or when it used to be one way, but now it is a different frequency, or not at all. This was ultimately not changed in the final version as the need to have a baseline estimation for hair removal/depilation outweighed variable responses.

### Example 5: Obstetrics-Intrauterine Growth Retardation

Survey Question: Sometimes babies are smaller than we expect. During this pregnancy did your baby have intrauterine growth retardation?

Finding: Respondents commented that they felt the first part of the question didn’t match the second part. They did not understand that the first sentence was supposed to be the definition of “intrauterine growth retardation.” Since the goal of the question was to assess the size of the baby, not whether the respondent understood the medical phrase “intrauterine growth retardation,” the question was modified to “Sometimes, babies are smaller than we expect. During this pregnancy, were any of your babies smaller than expected for their gestational age?”

### Example 6: Rotating Shift Work

Survey Question: A rotating shift is a work schedule where your hours change in a predictable way from day-to-day, week-to-week, or month-to-month. In the last month, did you work rotating shifts?

Findings: This is another example where a definition was provided, but respondents did not consistently understand the idea of a “rotating shift.” Rather than focusing on whether their hours changed in a “predictable” way, some simply answered about whether their hours changed at all in the last month. They included changes such as working overtime and having a job where the hours were different on different days (such as 9-7 on Mondays and 10-2 on Tuesdays). In future iterations of the survey, the research team removed this question and asked: Have you ever worked a job where you worked at least some time between midnight and 4:00 AM?

### Example 7: Other Comprehension Problems

Comprehension problems can arise from not understanding the words in the question and can also include not understanding abbreviations or symbols. While “lbs.” is the abbreviation for pounds, we had a respondent who did not know what it stood for. In several questions, the symbols “<” or “>” were used in the response options. While they are commonly used, several of the respondents were slowed down by seeing the symbol instead of the words and had to think about which symbol was “greater than” and which was “less than”. In the final version, we did not use any abbreviations or symbols.

### Recall and Answer Formation Problems

Cognitive testing also helps researchers understand whether respondents have the information needed to answer questions. Recall and answer barriers occur when respondents: cannot remember the information needed to answer a question, never had the information needed to answer a question, or have not thought about the information in the format that the question is asking.

#### Example 8: Ovulatory Infertility

Survey Question: Ovulatory infertility is infertility due to not making an egg every month. Have any of your female relatives been diagnosed with ovulatory infertility?

Findings: This question had two cognitive problems - a comprehension problem and a recall problem. Most respondents did not understand the term “ovulatory infertility” and whether it differed from any other kind of infertility. Additionally, none of the respondents knew whether any of their female relatives had it. Since we realized respondents would probably not be able to provide reliable information, we decided to delete the question from the final version. Instead of asking about ovulatory infertility, questions surrounding infertility were streamlined to ascertain if a participant tried to get pregnant for more than 6 months and then asking about possible causes such as fallopian tube complications or hormonal variations.

#### Example 9: Hormonal Contraception

Survey Question: Now think about all the different times you’ve taken hormonal contraception in your life. Counting only the times you were taking it, for how many years did you take hormonal contraception? (If you took it for less than 12 months, check the box below).

Findings: This question was difficult for those who used multiple forms of contraception and had started and stopped over time. Respondents had the information needed but had to figure out how to get it into the format required. For those who had used an injection, there was an additional layer of complexity stemming from the phrase “taking it” over a specific length of time. One respondent included the amount of time they thought the shot was supposed to last. Another responded as if it was a one-time occurrence and did not report it as any time. In the final version, an extra sentence was added to the question: “If you had an injection or implant, include the time it was effective.”

#### Example 10: Waist Size

Survey Question: What is your waist size? Please select if you are responding in inches or centimeters. If you are unsure please take your best guess.

Findings: Only one respondent answered this question, with the rest saying “don’t know” and refusing to guess. When interviewed, the respondents explicitly said that they don’t know their waist measurement. However, they offered to give something similar that they did know-their pants sizes. Since respondents were unable to answer this question, because they didn’t have the information required in the format required, we decided to delete this question from the final version.

#### Example 11: Fruit Juice Serving Sizes

Survey Question: A serving of fruit juice is 4 ounces (half a cup). In the past week, how many servings of fruit juice did you drink.

Findings: This question includes three different measurements that the respondent had to juggle in their mind when answering. While the concept of a “serving” is often used by dieticians and other health professionals, most people do not think in those terms. As such, this question ended up being very confusing to respondents. Even those who could clearly articulate what fruit juice they drank in the past week could not figure out how to answer this question accurately. Although this question is cognitively complex, it was kept in the survey. However, the time frame was modified from “in the past week” to “in an average day” to make it easier for respondents to answer.

### Usability Concerns

Cognitive testing may also provide information about how respondents navigate through a survey and if there are user-interface problems.

#### Example 12: Natural Body Hair

Survey Question: Please mark the image in each row that best corresponds with the amount of hair you have on that part of your body (Figure 2). Please think about your natural body state (without the use of hair removal procedures or treatments):

**Figure 1.**
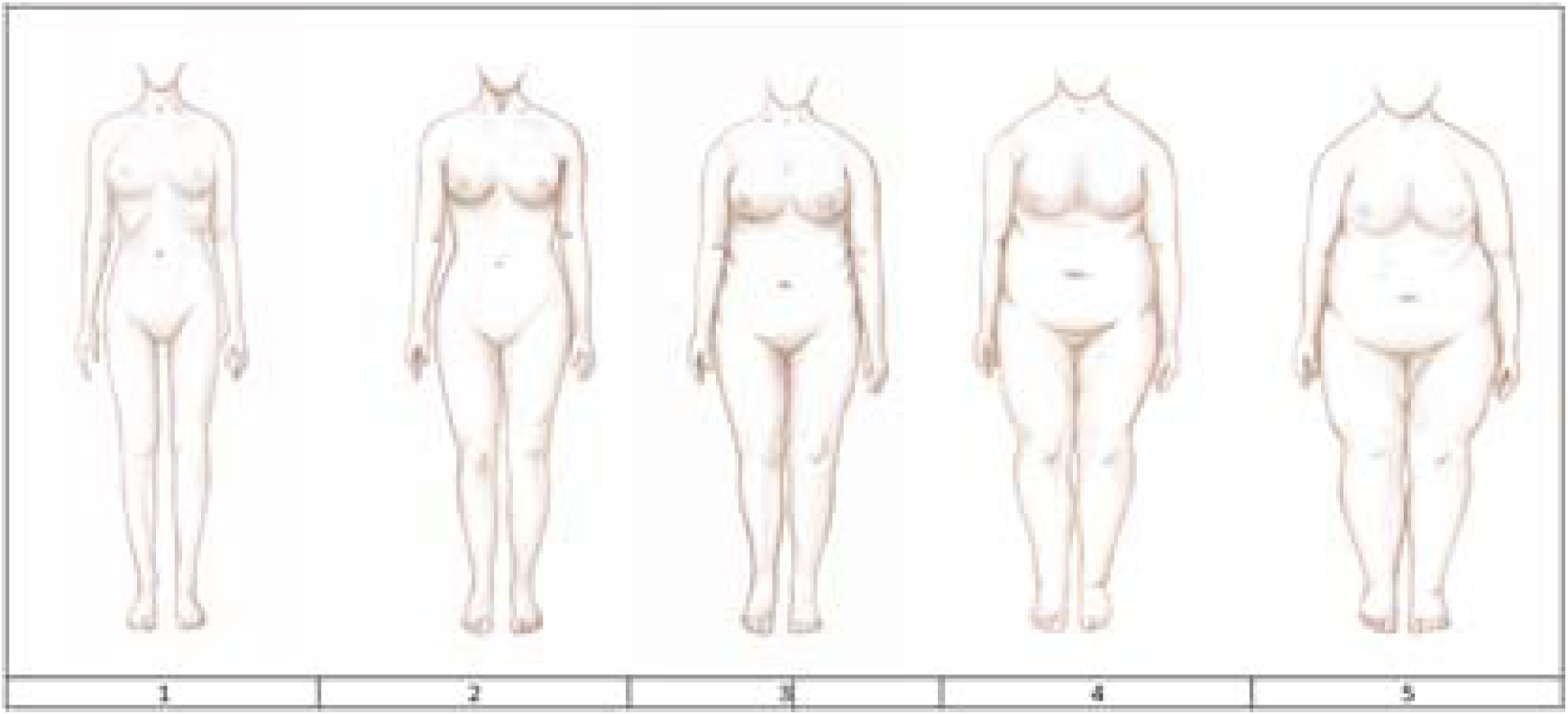
Anthropometries Image for Example 12.

**Figure 2.**
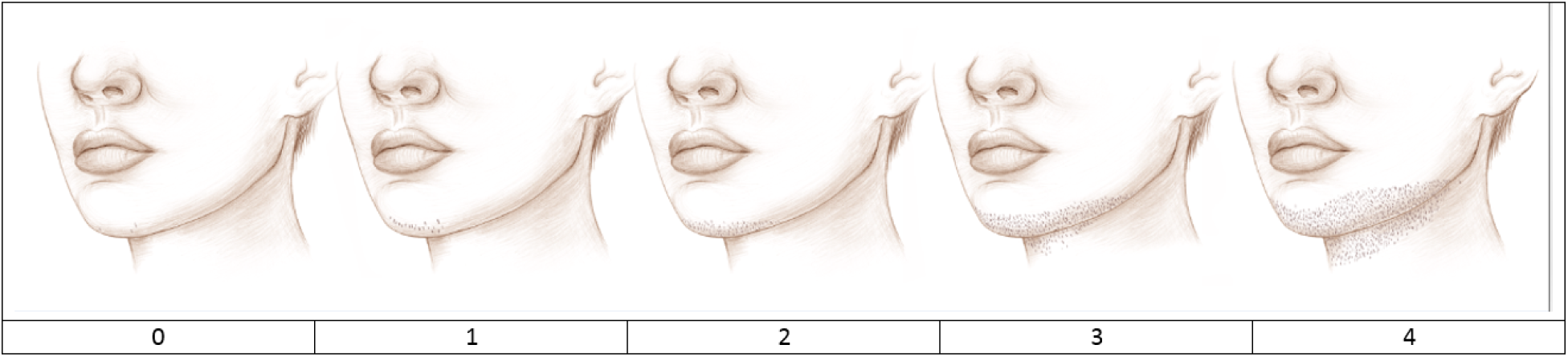
Chin Hair Hirsutism Rating Image for Example 13.

Findings: This question was followed by a series of pictures depicting hair on various body parts. Cognitive testing showed respondents understood what the phrase “natural body state” meant. As in the previous example, respondents had trouble answering the questions because they tried to actually click on or mark the image. Some respondents also said they had trouble seeing the distinctions between the options. In future versions, the format of the page was modified to make it easier for the respondents to understand they were supposed to select from answer choices below the image.

#### Example 13: Current Acne

Survey Question: In the last month or so, how would you currently rate your acne, also called pimples? (Figure 3).

**Figure 3.**
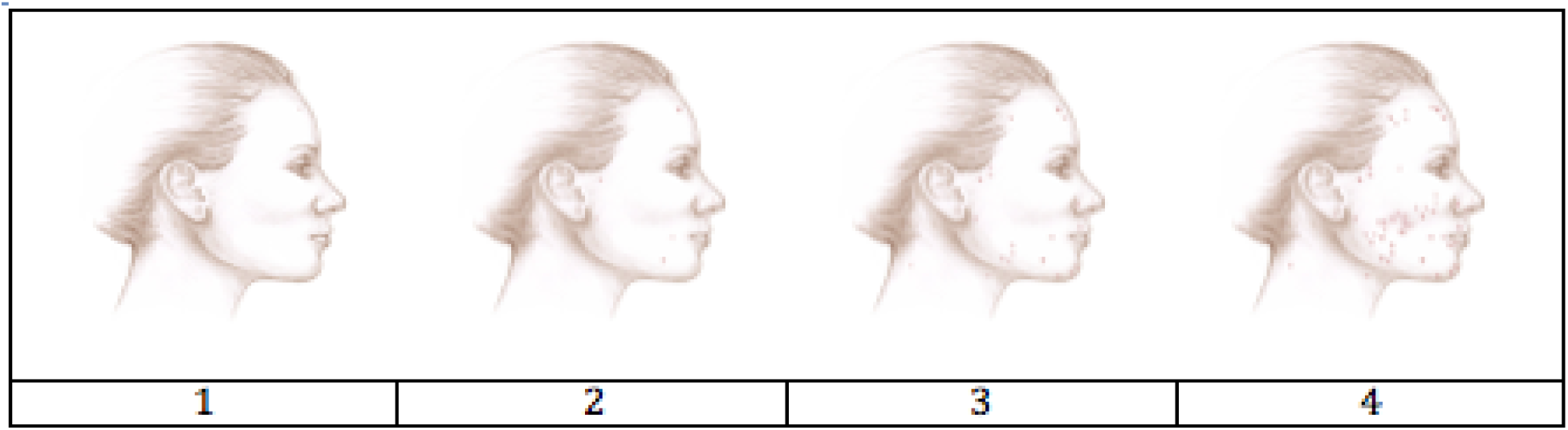
Facial Acne Rating Image for Example 14.

Findings: This question was followed by a series of answer choices with parenthetical descriptions of acne severity. Cognitive testing showed several respondents only looked at the pictures and did not read the descriptions in the answer choices. One respondent said it depends on the day/time of the month, and another asked whether or not to include acne on the back. Due to the text, parentheses, and pictures, the consultant suggested that the image was adding unnecessary complexity to the question. In the revised version, the image was removed and the question was updated to say: “Thinking about your face or back, how would you rate your acne? If your acne changes during your menstrual cycle, please think about your acne at its worst. [Response options] None or rare acne (none to a couple of pimples), Mild acne (4 or more pimples), Moderate acne (4 or more pimples that are red and irritated), Severe acne (4 or more pimples that are red, irritated, and have pus).”

#### Example 14: Anthropometrics

Survey Question: Now think about when you were 18 years old. Please mark the number displayed below the image which most looks like your body shape when you were 18 (Figure 1).

Findings: Respondents did not have any problems figuring out how to answer this as they knew which figure looked most like their body shape. The issue arose when they had to figure out how to mark their answers on the screen. Several respondents tried to click on the number below the picture, which did not work. They had to look further down the page to where the answer options were actually listed. After cognitive testing, the format of the page was modified in future versions to make it easier for the respondents to understand. The question was also changed to: “Which image above looks most like your body shape when you were 18?”

## Discussion

We present a cognitively tested survey instrument to ascertain menstrual cycle characteristics and androgen excess. To the best of our knowledge, no other instrument for self-assessment of menstrual cycle characteristics and androgen excess, inclusive of hirsutism, acne, and androgenic alopecia, exists for use in large population-based epidemiologic studies. Cognitive testing allowed for informed iterative survey design based on respondents’ comprehension of survey questions, including pictorial tools for image-assisted questions. Cognitive testing identified questions and concepts not easily comprehended, recalled, or with problematic response choices. Through the process of cognitive testing, we identified questions that would likely contribute to response errors and revised them.

For instance, example 1 demonstrated the importance of generating a clear definition for conditions pertinent to assessing PCOS. The creation of a definition that was simultaneously broad enough to capture the intended group and specific enough to allow for a prevalence assessment of PCOS had to be balanced. Similarly, example 3 which asks about menstrual cycle and definition of bleeding days demonstrates the importance of the location of the definitions. Knowing that respondents consistently read only to the question, the location of the spotting definition became critical to ensure participants were reporting the appropriate menstrual cycle length.

Additionally, expanding descriptions to capture the intention of the question was important. In example 9, identifying a timeframe for hormonal birth control usage helped elucidate the intention of the question which was to identify a duration of use by contraceptive formulation as injectables may have an efficacy of a month to several months. In contrast, we decided to retain questions such as example 4, which asks about frequency of hair removal. Despite issues quantifying frequency of hair removal amongst respondents, we felt the need to have a baseline estimation for hair removal/depilation to asses average depilation frequency in women and kept this question in.

### Implications of Cognitive Testing

When survey questions are not consistently understood and answered by respondents, there is a high likelihood of response errors and resulting biased findings. Cognitive testing is an evaluation tool that allows researchers to fix problems with survey questions before the survey is fielded. This evaluation serves two main functions: 1) it allows researchers to better understand if questions are working as intended and 2) it provides investigators insight into how respondents handle the cognitive steps involved in answering questions (which, if needed, provides insight into how a question might be improved). These cognitive steps include *comprehension* (were the questions consistently understood by respondents as intended by the researchers); *recall* (does the respondent have the information needed to answer the question); and *judgement and answer formation* (were respondents able to decide and provide an answer that accurately reflected what they wanted to say in the format the researchers required). Cognitive testing allows researchers to be more confident that survey respondents understand questions the way researchers intend. Its use in questions relating to critical outcomes of the study may prevent inadvertent errors in questioning which later need to be adjusted, resulting in missing data and/or improper answering of key questions.

The use of cognitive testing as a survey evaluation tool has become standard practice by survey researchers when creating a new survey or when using a survey originally created for a different population. It is likely that many current large-scale cohort studies involving surveys undergo some form of cognitive evaluation before they go into the field. However, surveys designed before the 1980s were probably not cognitively tested (Beatty & Willis, 2007) (Willis, 2004). Questions initially designed for the Nurses’ Health Study 2 (which began in 1989), were designed for health professionals, and assume the respondents have a high educational level and medical knowledge, an assumption that might be incorrect for the general population (Bao et al., 2016). For example, the rotating shift may have been widely understood when it was used in the Nurses’ Health Study 2 (Vetter et al., 2016). However, it was poorly understood by several of our respondents during cognitive testing (see example 6). Even for studies that are focused on the same population, researchers need to consider the changes in population over time. For example, a study conducted using race/ethnicity questions from the 1900s will not be reflective of the demographics in the United States in 2020s (*What Census Calls Us: A Historical Timeline*, 2015). Therefore, researchers must consider the implications of using survey questions that were created in a different time period. Similarly, social and temporal factors play a major role in determining the boundary conditions in which a cognitively tested tool remains functional.

The questionnaire and pictorial tools were designed from our review of the strengths and limitations of questionnaires from large cohort studies, such as the Nurses’ Health Study (NHS2) and the Framingham Heart Study (FHS). While, ascertainment of PCOS was limited in comparison to the survey we developed, they served as an important foundation for identifying the current gap in ascertaining information about self-reported androgen excess in women. Examples 12 and 14 demonstrate that image assisted questions were generally well comprehended.

Limitations of this survey instrument include the need to validate self-reported menstrual cycle characteristics and androgen excess against a gold standard. This gold standard may be an in-person interview or a medical record based clinical determination to compare with the self-reported responses. Validation of self-reported features against a gold standard such as medical record validation of PCOS diagnosis, reproductive health conditions, and androgen excess will be undertaken in future studies. Although cognitive testing is often conducted with a small number of respondents, researchers have found that important information can be gleaned about potential problems using a small sample size, especially if several respondents encounter the same issues or misunderstand the same things. Since the purpose of cognitive testing is to evaluate the survey instrument, in order to be most effective, respondents should come from the population for which the survey is intended as this study has done.

This survey instrument has several strengths. We ask about components critical to the diagnosis of PCOS, including androgen excess and menstrual irregularity. We are able to define PCOS using a composite of symptoms based on clinical diagnosis rather than using a yes/no response. This is key to ascertain those with milder forms of the disease in the general population. Using this new survey instrument, we can assign a PCOS state beyond the binary answers to menstrual irregularity questions. Furthermore, this survey instrument improves upon existing cohorts lack of specificity in the question stems asked.

This survey instrument was designed for those with an 8^th^-grade level English language proficiency and can be easily shared to facilitate harmonization across cohorts. We have shared this entire instrument in a genomic study, Genes for Good, to ascertain phenotypes of menstrual irregularity and androgen excess (*Genes for Good: For Researchers*, 2019). This tool is complementary and additional to phenotyping suggested by the PhenX ToolKit, whose questions are limited to androgen excess in males only at this time (*Study For Future Families*, 2020) (Swan et al., 2003). Other uses include employment in areas of the world that have not had appropriate ascertainment or studies related to menstrual cycle research. The pictorial tool has already been shared with researchers leading the Boston University Pregnancy Study Online (PRESTO) to evaluate hirsutism within a cohort of North American Women attempting pregnancy (Willis et al., 2020).

In summary, this instrument includes targeted questions and images for anthropometrics and androgen excess, including acne, hirsutism, and alopecia. Its overarching goal is to accurately ascertain cases of PCOS, some of whom may be misclassified as non-diseased.

## Conclusion

We designed and cognitively tested a questionnaire and pictorial tools to assess the menstrual cycle and self-reported androgen excess in a diverse population. This tool can be shared across studies to facilitate ascertainment of PCOS and phenotyping of menstrual disease. Given the long-term health risks for women with PCOS, it is imperative to have tools to identify affected women for possible interventions.

## Data Availability

Harvard Dataverse: Mahalingaiah, Shruthi, 2020, “The Ovulation and Menstruation Study Report on Cognitive Testing of Survey Instrument, July 5, 2017”, https://doi.org/10.7910/DVN/J0CL9O, Harvard Dataverse, V1

https://dataverse.harvard.edu/dataset.xhtml?persistentId=doi:10.7910/DVN/J0CL9O

## Data Sharing Statement

All data generated or analyzed during this study are included in this published article or in the data repositories listed in References.

